# A Landscape Analysis of the Recent Rise of Healthcare Data Science Degree Programs

**DOI:** 10.1101/2022.12.14.22283366

**Authors:** Aman Bhandari, Austin Cohen, Maxwell Fung, Trishan Panch, Alex Aronov, Nicholas Michaud

## Abstract

There has been an explosion in formal data science and healthcare data science programs, degrees and tracks at the university level over the last five years, with over 100 universities currently offering formal data science degrees. We conducted a landscape analysis of healthcare-related data science programs in the United States to characterize these new academic degree offerings, identifying 29 healthcare focused data science programs at four-year colleges and universities in the United States. Programs were analyzed across a range of features, providing the first view into what constitutes “healthcare data science” education.

## INTRODUCTION

Over the last decade, the combination of increasing amounts of data, advances in computing and developments in advanced statistical techniques has fueled the rapid rise of data science (here we use data science to also capture the popular uses of the terms AI and machine learning). In turn, there has been rapid growth in formal data science programs, degrees, and tracks at the university level over the last five years, including many healthcare specific programs. This phenomenon is not isolated to the United States. The 2021 Artificial Intelligence Index Report [1] notes “Since Canada published the world’s first national AI strategy in 2017, more than 30 other countries and regions have published similar documents as of December 2020.”, and more recently Fatima et al. [2] found over 40 countries have a national AI plan. China, for example, has created an “AI Innovation Action Plan for College and Universities” where 283 universities are offering licensed data science programs, and recently “35 [Chinese] universities received approval to establish the four-year undergraduate AI-related majors amid the country’s drive to build a strong AI talent pool” [3]. Globally there has been a significant investment by universities in this new official degree and associated course offerings at both the graduate and undergraduate levels.

While these programs have already begun to train the next generation of healthcare data researchers in the US and abroad, little work has been done to understand their educational content. The absence of a standardized set of classes to take and subjects to learn has the potential to result in learning gaps that could have real-world implications for graduates of these new programs.

In an effort to understand the current state of this rapidly expanding field, we catalogued every four-year college and university-based healthcare data science program in the country, recording key aspects of each program. We then summarized the data to understand what is present, and what is lacking, across these programs. Our analysis gives the first holistic view of the educational makeup of healthcare data science degree programs, and constitutes an initial effort at understanding their commonalities and differences.

## METHODS

Given the recent emergence of the healthcare data science field, there is no standard way to define which academic programs are “healthcare data science” programs. As such, we propose a set of three simple criteria for defining such programs:

1. The program offered a certificate, track, or degree in 2020 at a four-year college or university
2. The program title had “Data Science” in its name
3. Either:
  - the program is housed in a Biostatistics department, School of Medicine, Public Health or other Allied School of Health, or
  - the program title has “bio” or “health” in its name

These criteria result in a set of programs that are self-described as “data science” programs. Note that rather than attempt to define and include other similar types of programs in our data, we purposefully excluded programs that did not use the term “data science” in their title but may have been related, with titles such as: “informatics” or “business analytics”.

Applying the above criteria to a manually collected list of data science programs resulted in a set of 29 healthcare data science programs in the US. We then extracted course names and program descriptions for each identified program, resulting in information on 462 courses across the 29 programs. We constructed a dataset of these programs with the following features: *location, name of the program, type of program, department, program start date, program description text, and course titles*. Data was collected through each program’s website, and when information could not be located, the program’s administration was contacted to gather it directly.

Once collected, our analysis focused on summarizing the new data set. In addition to examining trends in when programs were created and what departments they are housed in, we conducted text analysis on course titles and program descriptions to infer broad themes, areas of focus, and gaps across healthcare data science programs. We categorized each course into a “field” by searching for keywords within that course’s title. For example, courses were categorized as “Statistics” if their title contained any of the words: “Statistics”, “Model”, “Regression”, “Multivariate”, “Analysis”, and “Probability”. ^1^ We also searched for keywords within program descriptions, providing a view into what themes programs use to define themselves.

All data used for the analyses in this paper is available in Supplemental Table 1.

## RESULTS

We found 29 healthcare data science programs in total. Four schools offered both a Masters and a Ph.D. program in the same department and one school offered a healthcare data science program in two separate departments. There were 23 Master’s programs, 7 PhD programs, and 3 bachelors programs (see Figure #1).

**Figure 1:**
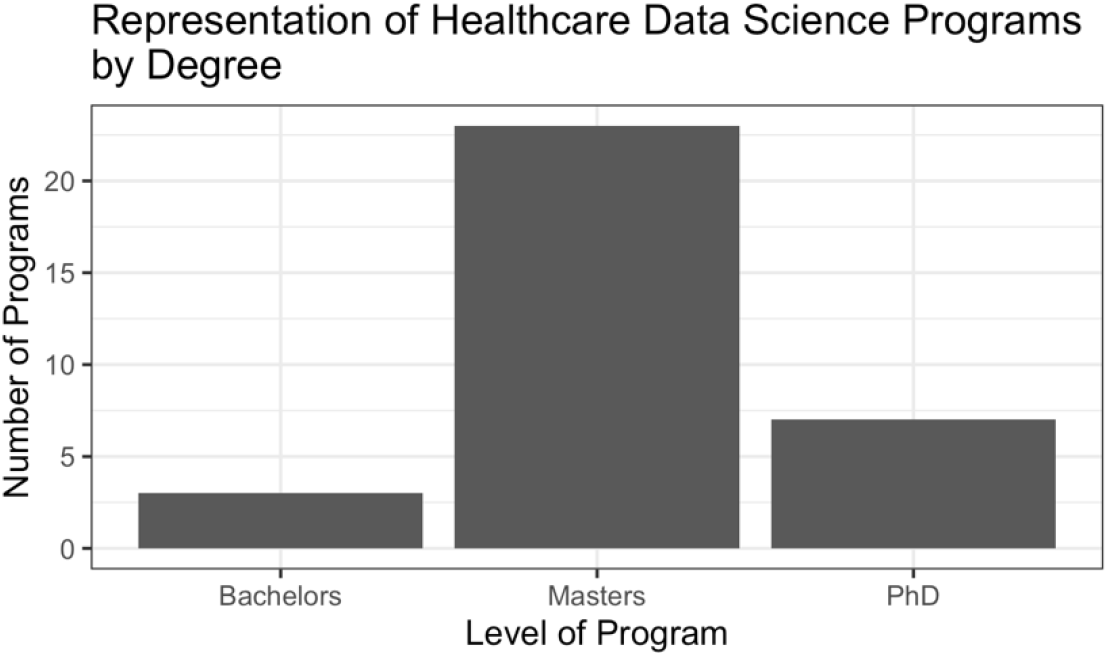
Counts of degree programs by degree type.

All of the healthcare data science programs we found were created within the last five years, highlighting the rapid expansion of this field. Of the programs that published a starting date, one was launched in 2016, eight in 2017, four in 2018, eight in 2019, and three in 2020. For the remaining five schools we were unable to get starting dates.

### Text Analysis of Program Descriptions and Course Titles

Program descriptions provide a broad overview of the content and focus of degree offerings, and can give insight into what skills and knowledge graduates are expected to carry into the workforce. Analysis of the text of the overall program descriptions revealed that about 75% of the schools mentioned statistics in their program description, 59% mentioned computer science and computation, and about 20% of the descriptions mentioned math or mathematics (all program descriptions are available on request to the author). Less mentioned topics included “programming”, which was mentioned 17% of the time, “engineering” and “machine learning” were each mentioned 10% of the time, and “AI/artificial intelligence” was mentioned 7% of the time. None of the program descriptions included mention of “ethics” or “bias”, and only one mentioned “inequality”, “inequity”, “disparities”, or “social justice”.

To understand more specific patterns in curricula across healthcare Data Science programs, we also analyzed the titles of required and optional courses for each program. We found 462 unique course titles across all 29 programs. Figure 2 displays the percent of programs with at least one listed course in each of our specified subject areas. Notably, over 93% of programs offered courses explicitly related to computer science, and over 89% offered statistics courses. On the other hand, only 17% of programs offered courses that explicitly addressed ethics, equality, or responsibility. This lack of ethical coursework mirrors the omission of ethics and equality from every data science program description in our dataset. For course titles which included the programming language they were being taught: 50% were in SAS and 38% were in R, which appeared to be favored over the 8% of classes in Python and 4% in SPSS.

**Figure 2:**
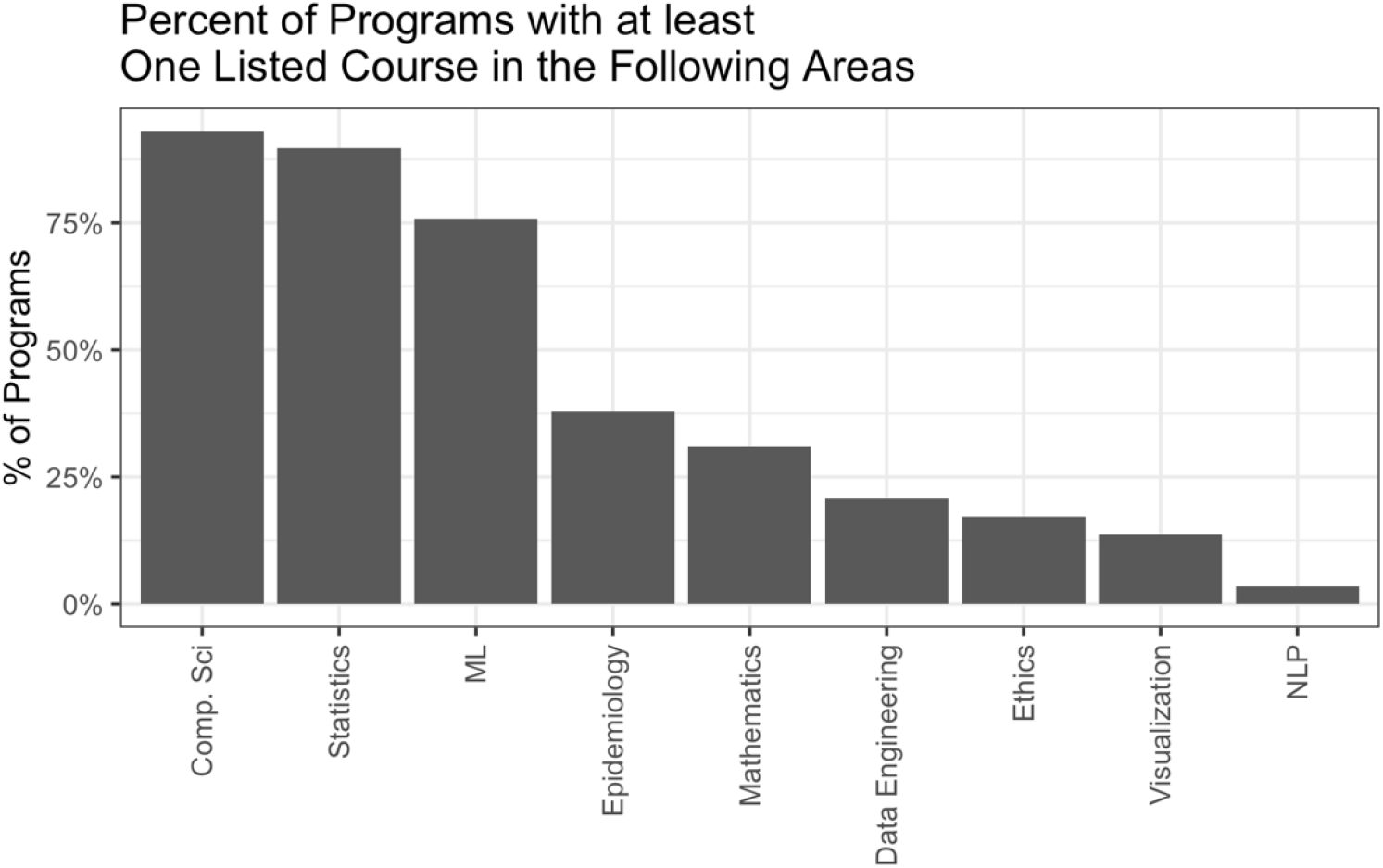
The percent of healthcare data science degree programs with at least one course in a given subject area.

### Trends in Biostatistics-Focused Data Science Programs

One notable category of programs in our data is data science programs that are biostatistics-focused. We defined this category in our dataset by searching for the term “biostatistics” in either the department name or the program name, identifying 14 biostats-focused programs. Many of these programs are former biostatistics degree programs that have changed their name to incorporate the term “data science”, as seen in Table 1. This shift towards incorporating data science into statistics education was highlighted in the 2016 Statistics and Biostatistics Department Chairs Workshop. The whitepaper for the workshop [4] notes “The rapid increase in the demand for data science training is profoundly affecting statistics education.”

**Table 1:** The first eight programs to change their name from biostatistics to include the term “data science”.

| University | Program Offering/Name Change | Date |
| --- | --- | --- |
| Tulane University | Global Biostatistics and Data Science | <u>2016</u> |
| Brown School of Public Health, Dept of Biostatistics | Health Data Science Track | 2016 |
| Harvard T.H. Chan School of Public Health | Master’s Program in Health Data Science | 2017 |
| The University of Texas Health Science Center at Houston | Department of Biostatistics and Data Science<br>- MPH/MS Concentration in Data Science | ~2016 |
| University of Wisconsin, Biostatistics and Medical Informatics | PhD programs in Biomedical Data Science | PhD new (Fall 2018) |
| Weill Cornell Medicine | MS in Biostatistics and Data Science | Fall 2017 |
| Stanford Medicine | Department of Biomedical Data Science<br>- Biomedical Informatics Graduate Certificate | Fall 2015 |
| Dartmouth Geisel School of Medicine | Department of Biomedical Data Science<br>-MS in Biostatistics | Fall 2018 |

To investigate what, if anything, differentiates these biostatistics focused programs from other healthcare data science programs, we compared course offerings between the two groups. Figure #3 compares rates of programming courses between biostatistics focused and non-biostatistics focused programs. Across all languages considered, courses involving programming languages were more common in the biostatics focused programs, indicating that such programs may have a more technical or more focused course load when compared to non-biostats programs.

**Figure 3:**
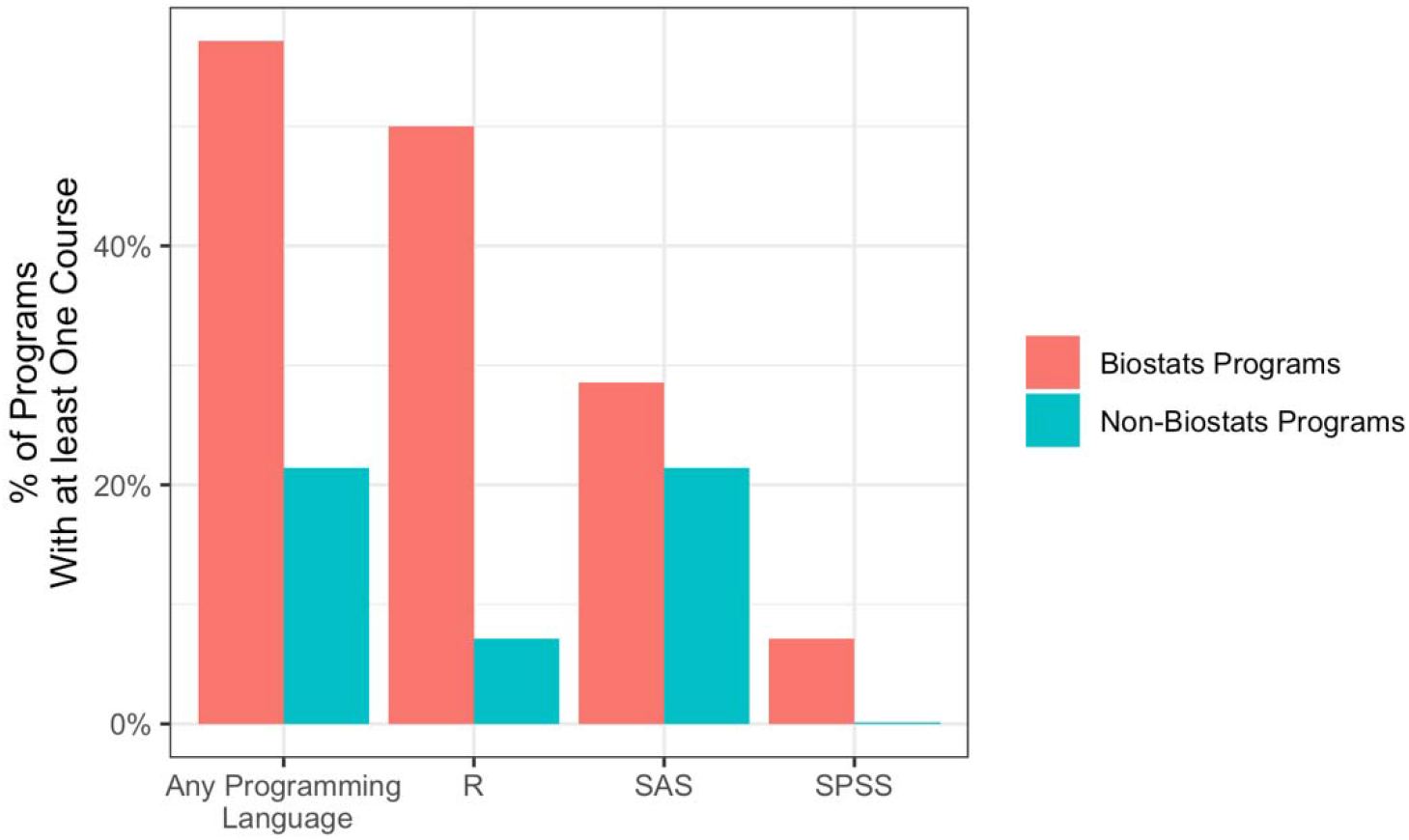
The percent of degree programs with at least one course title that mentions a programming language by name.

## DISCUSSION

With the launch of many campus-wide and department specific initiatives, data science is altering the educational landscape across all disciplines. Yet very little is known about these programs. In this analysis, we focused on healthcare data science programs in Biostatistics departments, Schools of Medicine, Public Health, and other Allied Schools of Health.

Based on our text analysis of program descriptions and course titles, we found three important teaching gaps that warrant further investigation. First, among the text of the program descriptions and course titles there was very little explicit mention of some key areas of data science knowledge, such as data engineering, machine learning, and natural language processing. And while we found some commonly used terms in program descriptions and course titles, there is a clear opportunity to work towards standardizing the meaning of healthcare data science and how it differs from other programs with names such as informatics or biostatistics. Lack of shared definitions and framework can lead to confusion, particularly given the rapid evolution in data science across a diversity of applications in healthcare. We recommend that programs should adopt a competency model which would standardize definitions, curriculum, and a minimum level of competency in key areas.

Second, for courses that specifically mentioned a programming language, SAS followed by R were the dominant offerings. However, a recent Kaggle survey [5] of almost 20,000 data scientists found the top programming languages in use to be Python, SQL, R and then Java, with R weakening significantly compared to prior years. This may indicate a significant gap between academia and industry.

Third, we found only 17% percent of all programs have a listed class related to ethics, inequality, responsibility, or bias. This mirrors a recent survey [6] that found only 15% of instructors/professors say they are teaching ethics for data science. While this set of topics might be covered elsewhere, the lack of dedicated classes is a major shortcoming in the offering of these programs, one that should be further investigated and addressed. This finding echoes the numerous calls for “algorithmic stewardship” [7] in the workforce, but in this context, should be applied to academic training as preparation. Texts such as “Ethics and Data Science” [8] could help to form the basis of data science ethics courses within the programs we found.

Our work has a few limitations. For one, our data set examines programs only within the United States. Additionally, we excluded programs that may have data science curriculum under names that don’t include the words “data science”. We note that the combined use of computational and statistical techniques has been a part of healthcare and life science education for at least the last two decades, and includes areas such as bioinformatics, computational chemistry, computational genomics, and clinical or health informatics. However, even with these academic disciplines, we believe “data science” itself as a new discipline has formally emerged among allied schools of health and it is an important phenomena worthy of further analysis.

In summary, there has been a fundamental shift across higher education, with newly launched programs on data science following the “irrational exuberance” [9] around the use of AI. These programs have an obligation to train the next generation of talent that is reflective of both the opportunities and issues in the workforce. Understanding the rise and current state of such programs is important for assessing gaps along with needs around standards as well as implications for training, funding and policy development of the next generation of talent.

## Data Availability

All data produced in the present study are included in the supplementary materials.

### Appendix

#### I: Keywords Used to Classify Courses

**Table 2:** Keywords used to categorize courses.

| Course Categorization | Keywords |
| --- | --- |
| Comp Sci. / Programming | comput*, CS, program, python, R, SAS, SPSS |
| Statistics | bayes, bootstrap, causal, data analysis, experiment, inference, linear, longitudinal, regression, sampling, stat*, survival |
| ML | ai, intelligence, learning, machine, ml |
| Epidemiology | epidem* |
| Mathematics | algebra, calculus, math, probability, real analysis, stochastic |
| Data Engineering | database, engineer* |
| Ethics | disparity, equality, equity, ethic*, justice, responsibility, responsible, |
| Visualization | display, graphical, visual |
| NLP | language, NLP |

The above table lists the different course categories and corresponding keywords that were used to create Figure #2. Keywords followed by an * would match any word that begins with those letters. Note that one class can fall into multiple categories.

#### II: Program Listings

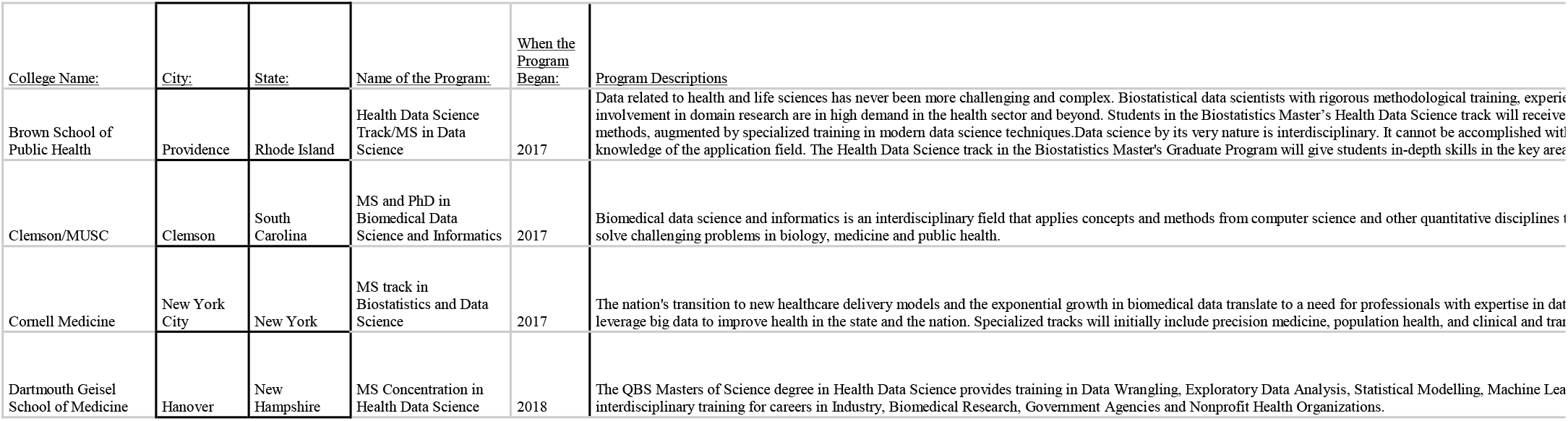
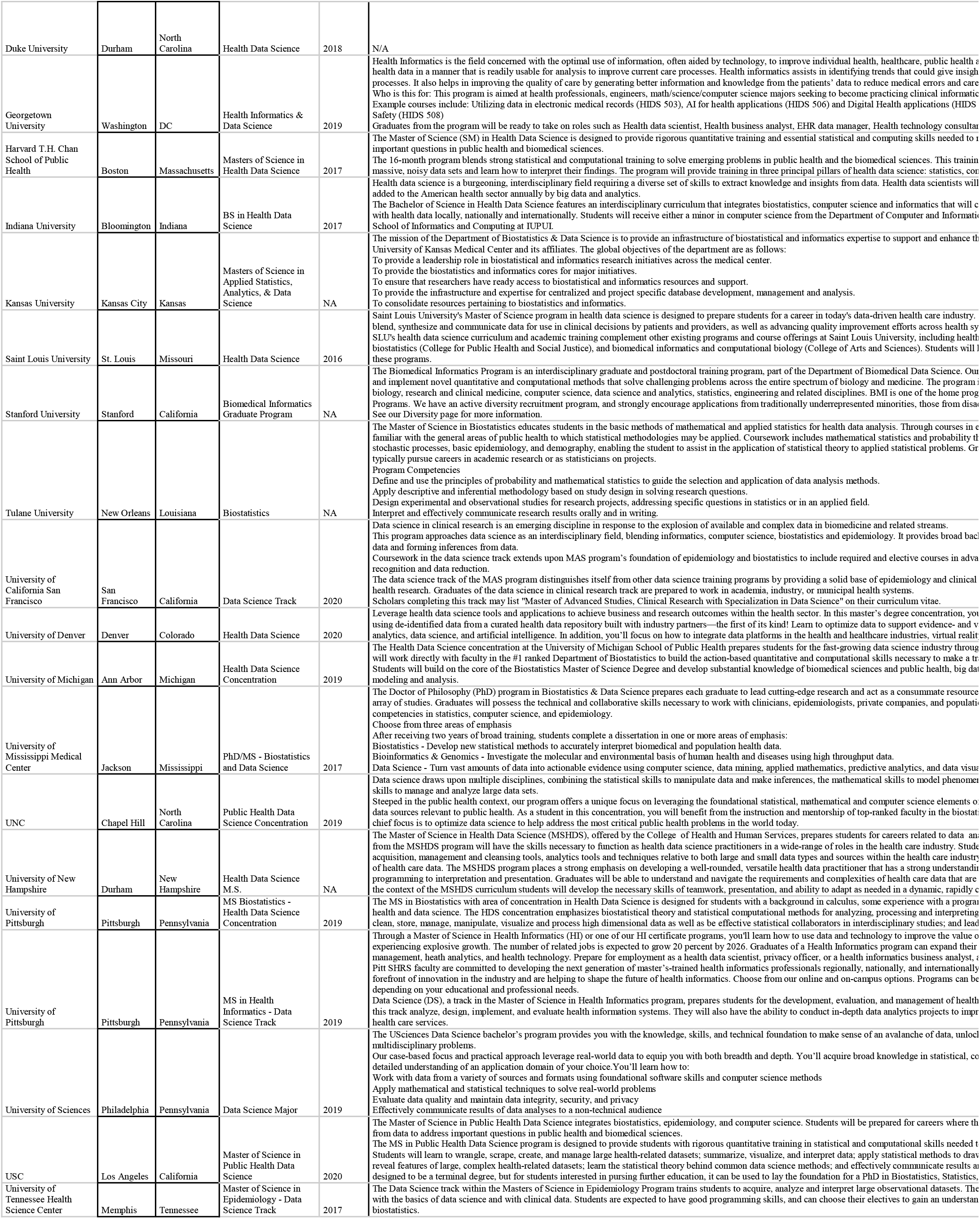
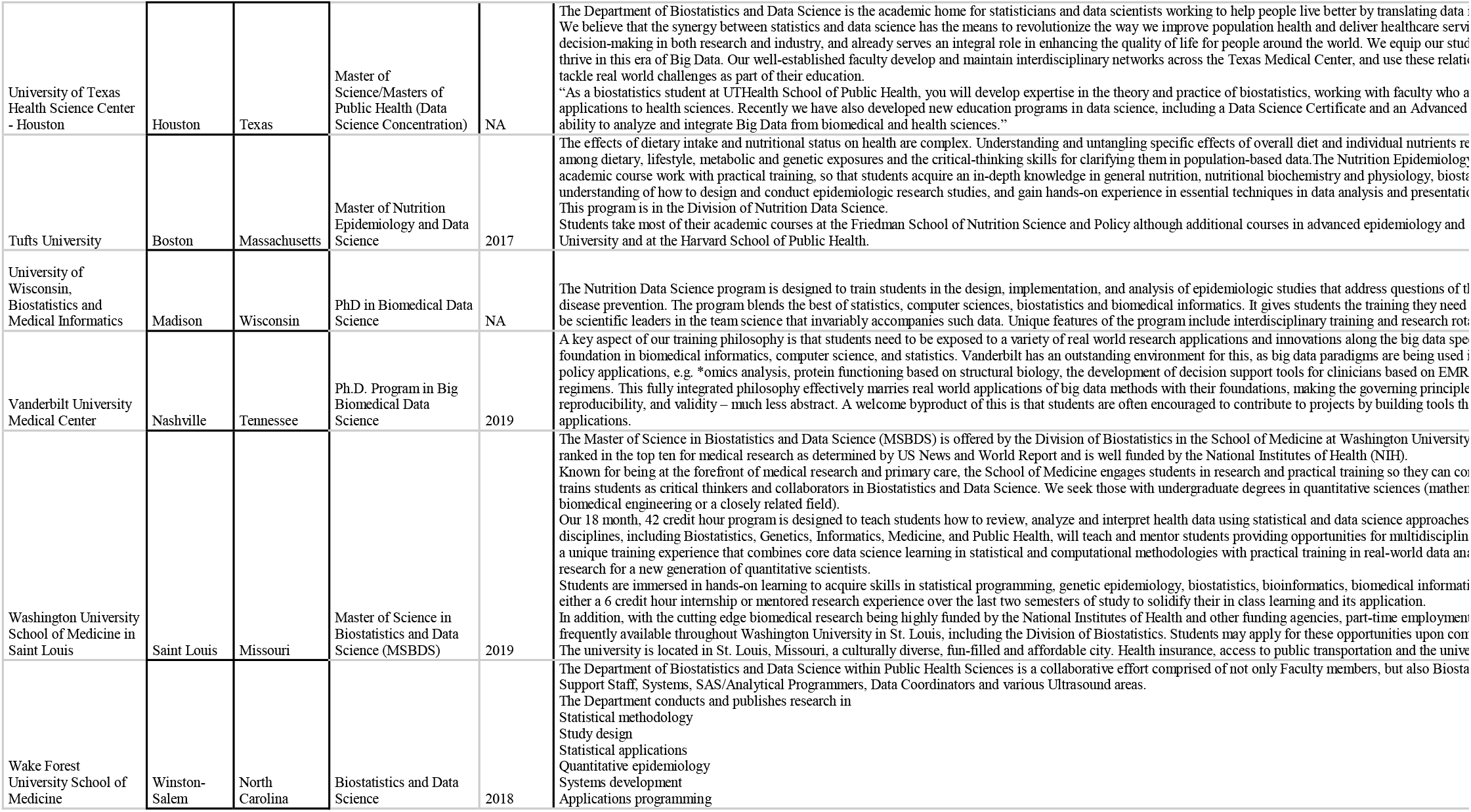

## Footnotes

1 For a complete list of course categorization keywords, see Appendix Section I.

